# Genomic Insights into the emergence of Carbapenem-Resistant *Salmonella* Typhi harboring *bla*_NDM-5_ in India

**DOI:** 10.1101/2025.05.20.25327816

**Authors:** T Tharani Priya, Jobin John Jacob, T Monisha Priya, Mahantesh, Bhavana, Suhani, Savitha Nagaraj, Jayanthi Savio, Priyadarshini A Padaki, J Sudarsana, Ashalatha Nair, Suganya Verma, Raman Gaikwad, Divya Joshi, Vasant C Nagvekar, Camilla Rodrigues, S Pavithra, B Santhosh K, Jacob John, Kamini Walia, Balaji Veeraraghavan

## Abstract

**Background:** The rising antimicrobial resistance in *Salmonella* Typhi presents a critical public health challenge by limiting effective treatment options. This highlights the importance of sustained genomic surveillance of the high-risk lineages and antimicrobial resistance determinants of *S*. Typhi to inform and support the implementation of typhoid vaccination programs, especially in endemic regions. In this context, we investigated a recent outbreak of carbapenem-resistant *Salmonella* Typhi in India, to elucidate the genomic epidemiology, resistance mechanisms, and evolution of drug resistance.

**Methods:** A total of 21 carbapenem-resistant *S*. Typhi isolates collected from multiple healthcare facilities were subjected to phenotypic and genotypic characterization. Short-read whole-genome sequencing (WGS) was performed to analyze the population structure, identify single nucleotide polymorphisms (SNP), and investigate the evolution of antimicrobial resistance. A maximum likelihood phylogenetic was constructed in the context of global isolates to infer the emergence and dissemination of carbapenem-resistant *S*. Typhi. Additionally, two isolates were subjected to long-read nanopore sequencing to characterize the plasmids and assess their circulation among other Enterobacterales.

**Results:** Antimicrobial susceptibility testing of isolates revealed resistance to ampicillin, ciprofloxacin, ceftriaxone, and carbapenem, while the isolates remained susceptible to chloramphenicol, cotrimoxazole, and azithromycin. The genomic analysis identified the presence of two plasmids: IncFIB(K) harboring *bla*_CTX-M-15_, *qnrS1*, *tetA*, *tetR,* and IncX3, carrying the *bla*_NDM-5_ gene. Phylogenetic analysis classified the isolates within a novel genotype, 4.3.1.1.1 belonging to genotype 4.3.1.1(H58 lineage I). The independent acquisition of resistance plasmids from other Enterobacterales in multiple *S*. Typhi genotypes highlights a significant public health concern.

**Conclusion:** Our findings emphasize the need for robust genomic surveillance to monitor antimicrobial resistance, trace the dissemination of high-risk *S*. Typhi lineages, and guide evidence-based antibiotic policies for the treatment of drug-resistant strains. Strengthening surveillance addresses travel-associated transmission and prevents the global spread of drug-resistant strains.

## Introduction

Enteric fever, primarily caused by *Salmonella enterica* serovars Typhi and Paratyphi A, remains a major public health challenge, particularly in low- and middle-income countries (LMICs) (Parry et al., 2002; Crump et al., 2015). This disease is primarily transmitted through the ingestion of food or water contaminated with fecal matter, potentially leading to severe complications, such as intestinal perforation, if untreated (Radhakrishnan et al., 2018; Crump, 2019). Recent estimates indicate a global burden of approximately 9.24 million enteric fever cases in 2021, with South Asia accounting for 62% of the total incidence. Within this region, India bears the highest burden, contributing 58% of global cases, followed by Pakistan and Bangladesh (Piovani et al., 2024). Notably, children under five years of age are disproportionately affected, experiencing a significant share of the disease’s morbidity and mortality, particularly in endemic areas (John et al., 2023).

The management of typhoid fever has historically relied on antimicrobial therapy; however, the emergence of antimicrobial-resistant (AMR) strains has significantly compromised treatment efficacy (Browne et al., 2024). In the late 20th century, multidrug-resistant (MDR) *S*. Typhi strains emerged, exhibiting resistance to first-line antibiotics such as ampicillin, chloramphenicol, and trimethoprim-sulfamethoxazole (Crump et al., 2015). This situation prompted a shift towards fluoroquinolones as the primary treatment option (Marchello et al., 2020). Unfortunately, the subsequent rise of fluoroquinolone non-susceptible (FQNS) strains necessitated the exploration of alternative therapies, including azithromycin and third-generation cephalosporins (Kirchhelle et al., 2019). In recent years, the emergence and spread of ceftriaxone-resistant *S.* Typhi strains in Pakistan and India has further complicated the treatment strategies. In Pakistan, the outbreak of extensively drug-resistant (XDR) *S*. Typhi, has spread across the country, with ∼5,274 cases reported by December 2018 (Akram et al., 2020). These XDR strains exhibit resistance to first-line antibiotics, fluoroquinolones, and third-generation cephalosporins, leaving azithromycin as the primary treatment options (Klemm et al., 2018, Akram et al., 2020). Similarly, ceftriaxone-resistant *S*. Typhi outbreaks have been documented in India, including in Mumbai and Vadodara, (Jacob et al., 2021, Thirumoorthy TP et al., 2025). Furthermore, azithromycin-resistant strains, driven by mutations in the *acrB* gene, threaten one of the few remaining oral treatment options, highlighting the urgent need for new therapeutic strategies (Carey et al., 2021).

Genomic surveillance is a key tool for monitoring AMR transmission and tracking outbreaks of *S.* Typhi (Baker et al., 2018). The GenoTyphi scheme provides a robust framework for classifying *S*. Typhi into four major lineages and over 75 genotypes, enabling the identification of distinct transmission pathways on a global scale (Wong et al., 2016; Dyson et al., 2021). Among these, haplotype 58 (H58, genotype 4.3.1) has become the dominant strain worldwide, primarily due to its enhanced transmissibility and MDR (Wong et al., 2015; Pragasam et al., 2020; Carey et al., 2024). Within this lineage, key subclades include 4.3.1.1, associated with MDR, 4.3.1.2, linked to FQNS, and 4.3.1.3, which predominates in Bangladesh, providing critical epidemiological insights into the evolution and spread of drug-resistant typhoid (Carey et al., 2023). Further the GenoTyphi framework has proven effective in managing outbreaks of drug-resistant *S*. Typhi. For instance, the XDR *S*. Typhi outbreak in Pakistan, attributed to subclade 4.3.1.1.P1, was tracked using this scheme (Klemm et al., 2018). Similarly, ceftriaxone-resistant *S*. Typhi outbreaks in India, including in Mumbai (subclade 4.3.1.2.1) and Vadodara (4.3.1.2.2) were effectively tracked and characterized (Jacob et al., 2021; Argimón et al., 2022; Thirumoorthy et al., 2025).

Recently, sporadic reports have highlighted the emergence of carbapenem-resistant *S*. Typhi (CRST) in Pakistan, raising serious concerns about the evolving landscape of antimicrobial resistance (AMR) in this pathogen (Ain et al., 2022; Nizamuddin et al., 2023). More alarmingly, similar cases have been reported in various cities across southern and western India, indicating a potential regional spread of this resistance profile in a region where typhoid fever remains endemic. Despite these developments, the genomic epidemiology and evolutionary trajectory of CRST strains in India remain poorly understood. In this study, we aimed to investigate the genomic characteristics and evolutionary dynamics of CRST isolates in India, with particular focus on their rising prevalence and the role of plasmids in mediating resistance. A deeper understanding of these factors is critical to inform public health strategies and guide targeted interventions to curb the spread of CRST in endemic settings.

## Methods

### Study Settings

A total of 22 *Salmonella* Typhi isolates were obtained between August 2024 and March 2025 from enteric fever patients treated at hospitals in various cities in South and West India and from individuals with recent travel exposure to these regions. Among the isolates, ten originated from St. John’s Medical College Hospital, Bengaluru, four from Fortis Hospital, Bengaluru, two from Baby Memorial Hospital, Kozhikode and one each from Indira Gandhi Institute of Child Health, Bengaluru, Agilus Diagnostics Lab, Mumbai, KIMS Health Hospital, Trivandrum, P.D. Hinduja Hospital and Medical Research Centre, Mumbai, Christian Medical College, Chittoor campus, Andhra Pradesh and Sahyadri Speciality Labs, Pune, Maharashtra. Participating institutions flagged these isolates due to atypical antimicrobial resistance profiles detected via phenotypic susceptibility testing and/or automated VITEK 2 systems. Detailed patient demographics, clinical affiliations, and travel histories are provided in Table S1. For in-depth analysis of resistance mechanisms, all isolates were transferred to the Department of Clinical Microbiology at Christian Medical College (CMC), Vellore, for whole-genome sequencing and genomic characterization.

### Bacterial isolates and Phenotypic testing

The isolates, once received at CMC Vellore, was cultured on blood and MacConkey agar plates to ensure purity. The isolates were confirmed *S*. Typhi by conventional biochemical tests, serotyping (Kauffmann-White scheme), and qPCR (Nair et al., 2019). Antimicrobial susceptibility (AST) of the study isolates were evaluated by Kirby-Bauer disc diffusion technique, with inhibition zone measurements and interpretations adhering to the Clinical and Laboratory Standards Institute (CLSI, 2024) criteria. The tested antibiotics included ampicillin (10 µg), chloramphenicol (30 µg), trimethoprim/sulfamethoxazole (1.25/23.75 µg), ciprofloxacin (5 µg), pefloxacin (5 µg), ceftriaxone (30 µg), cefixime (5 µg), azithromycin (15 µg), meropenem (10 µg), and ertapenem (10 µg). To complement these findings, the broth microdilution (BMD) method was employed to assess the minimum inhibitory concentrations (MICs) of additional antibiotics, namely cefepime, aztreonam, piperacillin-tazobactam, ceftazidime-avibactam, aztreonam-avibactam, and colistin. Colistin MIC was interpreted according to EUCAST guidelines (EUCAST 2024).

### DNA extraction and whole genome sequencing (WGS)

Genomic DNA was isolated from samples using the QIAamp® Mini Kit (250) (QIAGEN, Hilden, Germany) according to the manufacturer’s protocol. DNA purity and concentration were quantified using a Nanodrop One spectrophotometer (Thermo Fisher Scientific, Waltham, USA) and a Qubit Fluorometer with the dsDNA HS Assay Kit (Life Technologies, Carlsbad, USA). The presence of carbapenemase and ESBL genes were identified by multiplex PCR with an in-house gene panel, adapted from the methodology described by (Poirel et al., 2011).

For short-read sequencing, DNA was fragmented, and paired-end libraries were prepared using the Illumina Nextera DNA Flex Library Kit and Nextera DNA CD Indexes (Illumina, Massachusetts, USA). Equimolar library pools were sequenced on the Illumina NovaSeq 6000 platform (available at Unipath Specialty Laboratory Limited, Ahmedabad, India), generating 2×150 bp paired-end reads. All steps, including tagmentation, library amplification, and purification, were performed as specified by the manufacturer.

To complement the short-read data, long-read sequencing was performed on two isolates using Oxford Nanopore Technology (ONT). For each sample, approximately 200 ng of DNA was processed with the Nanopore Rapid Barcoding Kit 96 V14 (SQK-RBK114.96; Oxford Nanopore Technologies, Oxford, UK) following the manufacturer’s protocol. Sequencing was carried out on a PromethION P2 Solo platform with real-time base-calling enabled during the run. Basecalling of POD5 files was executed using Dorado v0.8.3 (Oxford Nanopore Technologies) with the dna_r10.4.1_e8.2_400bps_sup@v5.0.0 model to ensure high-accuracy sequence reconstruction.

### Quality Control, Assembly and Annotation

Quality assessment of Illumina reads was performed using FastQC v0.12.1 followed by adapter and index trimming with Trimmomatic v0.39. Contaminant screening and filtering were executed with Kraken v1.1.1 (https://github.com/DerrickWood/kraken), followed by sequence coverage analysis. High-quality reads (Phred score >30) were assembled into draft genomes using SKESA (https://github.com/ncbi/SKESA). For hybrid assembly, the Hybracter pipeline v0.8.0 (https://github.com/gbouras13/hybracter) was employed to process Oxford Nanopore (ONT) long reads. This workflow first improved the read quality with Filtlong, assembled long reads de novo using Flye, and polished the assemblies iteratively with Medaka. Further refinement was achieved by polishing Illumina short reads via Polypolish and PyPolca. Final genome completeness and accuracy were evaluated using QUAST v5.2.0. Genome annotations were performed using Bakta v1.10.1 (https://github.com/oschwengers/bakta) Unless specified, default parameters were applied throughout all analytical steps.

### Comparative genome analysis

Draft genome assemblies were analyzed with SeqSero v2.0 (https://github.com/denglab/SeqSero2) to verify the antigenic composition of the serotype. In silico multilocus sequence typing (MLST) was performed on all isolates using the pipeline provided by the Center for Genomic Epidemiology (https://cge.food.dtu.dk/services/MLST/). Antimicrobial resistance (AMR) genes were identified using NCBI AMRFinderPlus v4.0.3 (https://github.com/ncbi/amr). Plasmid content was determined by querying genome sequences against the PlasmidFinder database (https://cge.food.dtu.dk/services/PlasmidFinder/). Plasmids were compared using the Basic Local Alignment Search Tool (BLAST), and circular maps were prepared using the Proksee server (https://proksee.ca/) (Grant et al., 2023).

### Genotyping and Phylogeny

The isolates were assigned to previously defined genotypes using the GenoTyphi pipeline (available: https://github.com/katholt/genotyphi). Unique single nucleotide polymorphisms (SNPs) characterizing the novel sub-lineage were and subsequently incorporated into the GenoTyphi framework to track the outbreak in future investigations.

For phylogenetic analysis, genome assemblies of *S*. Typhi (n=429) representing all major genotypes were obtained from the curated global collection provided by the Global Typhoid Genomics Consortium (https://bridges.monash.edu/articles/dataset/Global_Typhoid_Genomics_Consortium_2022_-_Genome_Assemblies/21431883) (Carey ME et al., 2023), with data sourced from NCBI (Table S2). The sequencing reads were aligned to the reference genome of *S*. Typhi□CT18 (GenBank: AL513382.1) using Snippy v4.6.0 (https://github.com/tseemann/snippy). The resulting full alignment was processed using Gubbins v3.3.3 (https://github.com/nickjcroucher/gubbins) to remove recombination sites (Croucher et al., 2015). SNPs were extracted using SNP-sites v2.5.1 (https://sanger-pathogens.github.io/snp-sites/) (Page et al., 2016). A maximum-likelihood phylogeny was then reconstructed from the filtered alignment using IQ-TREE under the TVM+F+ASC+G4 model with 1000 bootstraps. The resulting phylogenetic tree was visualized and annotated using the Interactive Tree of Life software (iTOL v.5) (Letunic and Bork, 2021).

## Results

### Identification of CRST isolates and resistance profile

A total of 22 non-duplicated *S.* Typhi isolates were included in this study, collected from patients diagnosed with typhoid fever across multiple healthcare institutions in southern and western India, between 2024 and 2025. All isolates were confirmed as *S*. Typhi through standard biochemical tests, conventional serotyping methods, and qPCR. AST by disk diffusion revealed a consistent resistance profile across all *Salmonella* Typhi isolates except one, with resistance to ampicillin, ciprofloxacin, ceftriaxone, and carbapenems. One isolate was resistant to ampicillin, ciprofloxacin, and ceftriaxone but susceptible to carbapenems. (Table 1). Conversely, all isolates remained susceptible to chloramphenicol, trimethoprim-sulfamethoxazole, and azithromycin.

**Table 1:**
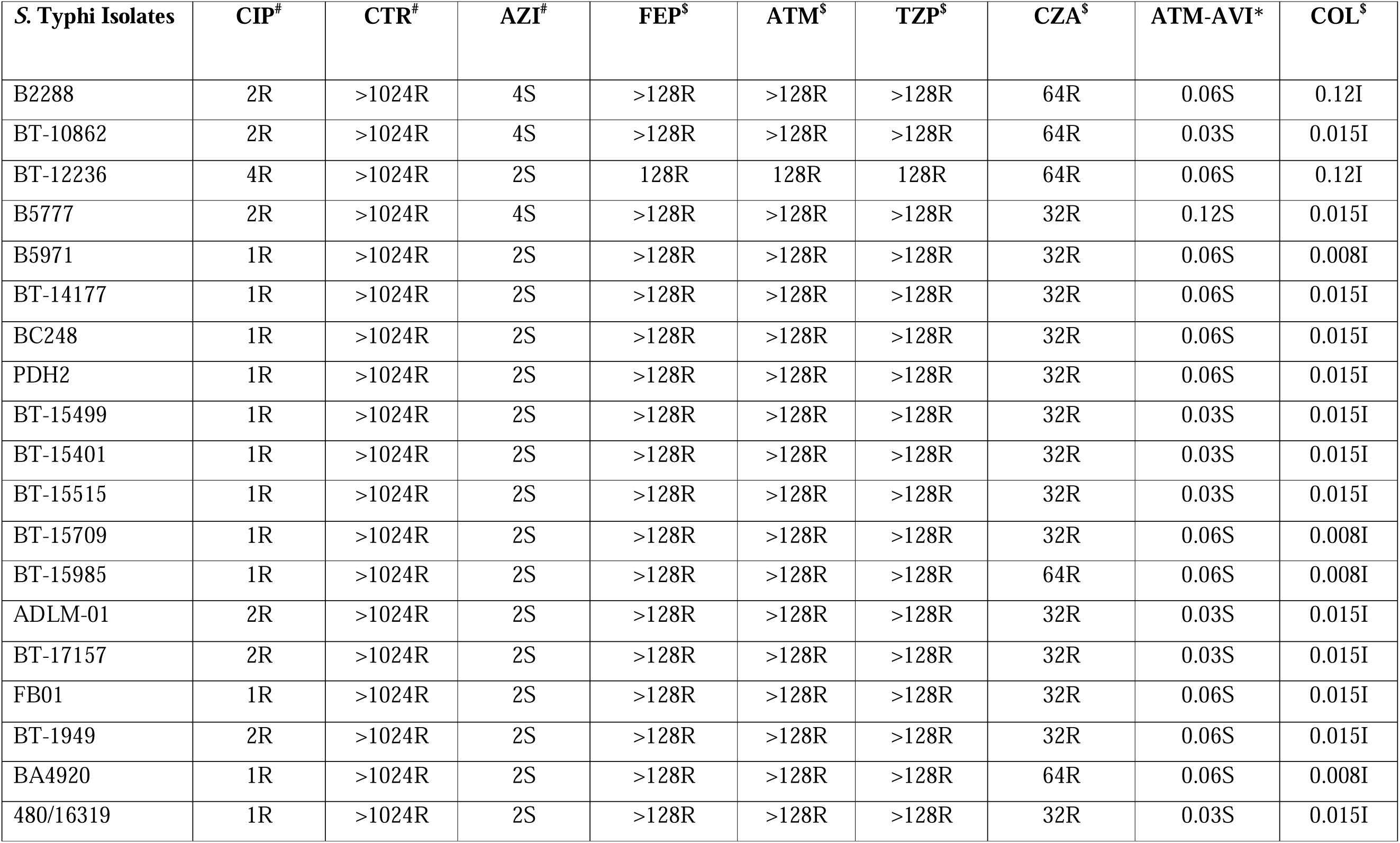
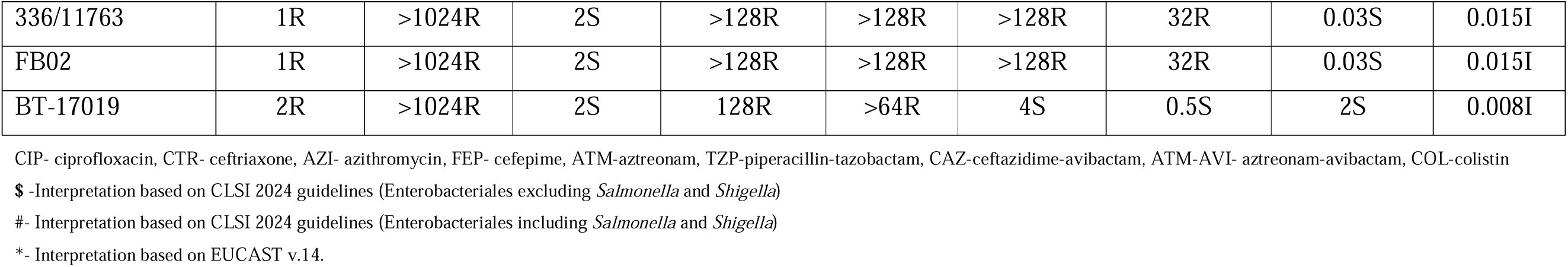
Antimicrobial susceptibility profile and minimum inhibitory concentration (MIC) in µg/mL of different antibiotics against *S*. Typhi isolates.

MIC testing further confirmed high levels of resistance, with MIC values of 2□µg/mL for ciprofloxacin, >1024□µg/mL for ceftriaxone, >32□µg/mL for ertapenem, and >16□µg/mL for meropenem. In contrast, chloramphenicol (2□µg/mL), trimethoprim-sulfamethoxazole (0.06–0.12□µg/mL), and azithromycin (2–4□µg/mL) retained susceptibility against all isolates. To assess the efficacy of β-lactam/β-lactamase inhibitor (BL-BLI) combinations against CRST, additional MIC testing was performed. The MIC ranges were as follows: cefepime and aztreonam (>128□µg/mL), piperacillin-tazobactam (>128□µg/mL), ceftazidime-avibactam (64–128□µg/mL), aztreonam-avibactam (0.03–0.06□µg/mL), and colistin (0.012–0.12□µg/mL). The multiplex PCR analysis confirmed the presence of bothlJ*bla*_NDM_□and□*bla*_CTX-M_□genes in all isolates, indicating the co-occurrence of carbapenemase and ESBL determinants among the CRST strains.

### Genotyping and comparative genome analysis

The□*S*. Typhi□genomes (n = 22) were characterized using the GenoTyphi genotyping scheme. All isolates were identified as belonging to the H58 haplotype (genotype 4.3.1), specifically falling within the 4.3.1.1 genotype (H58 Lineage I)(Figure S1). Based on their shared genomic features and epidemiological significance related to carbapenem resistance, these isolates have been assigned to a novel sub-genotype within 4.3.1.1, designated as 4.3.1.1.1.

AMR gene profiling identified the presence of *bla*_NDM-5_, *bla*_CTX-M-15_, *qnrS*, and *tetA* among the isolates. In addition, resistance-associated point mutation analysis revealed an S83Y substitution in *gyrA* within the quinolone resistance-determining region (QRDR). The presence of *bla*_NDM-5_ correlates with resistance to carbapenems, *bla*_CTX-M-15_□confers resistance to third-generation cephalosporins (3GCs),□*tetA*□is linked to tetracycline resistance, while both the□S83Y mutation in□*gyrA*□andlJ*qnrS*□contribute to fluoroquinolone resistance. Among the two plasmids identified IncFIB(K) carried *bla*_CTX-M-15,_ *qnrS*, and *tetA,* while *bla*_NDM-5_ was harboured by IncX3 plasmid.

### Population structure of CRST isolates from India

A core genome SNP-based phylogenetic analysis, incorporating the 21 CRST study isolates and 312 global reference isolates, revealed that the CRST isolates formed a distinct subclade within the H58 lineage I (genotype 4.3.1.1) (Figure 1). This CRST subclade was separated from its parent clade by seven unique SNPs, defining its distinctiveness (Table S3). Notably, the CRST isolates demonstrated the closest genetic relatedness (9 SNPs difference) to a previously sequenced isolate from Anantapur, Andhra Pradesh, India (ERR5200999). The most closely related isolate from one of the same study locations (Bangalore) was strain ERR5201244, which differed by 12 SNPs and was sequenced in 2018 as part of the Surveillance of Enteric Fever study. Isolates closely related to the CRST clone, but lacking resistance plasmids, have been circulating in India since at least 2018. This suggests that the CRST clone likely emerged from these endemic lineages through the recent acquisition of resistance plasmids. Furthermore, the CRST isolates sequenced in this study were phylogenetically distinct from both the previously reported CRST isolate from Pakistan (SRR22801806; 26 SNPs difference) and the ceftriaxone-resistant outbreak isolates recently identified in Gujarat, India (25 SNPs difference).

**Figure 1:**
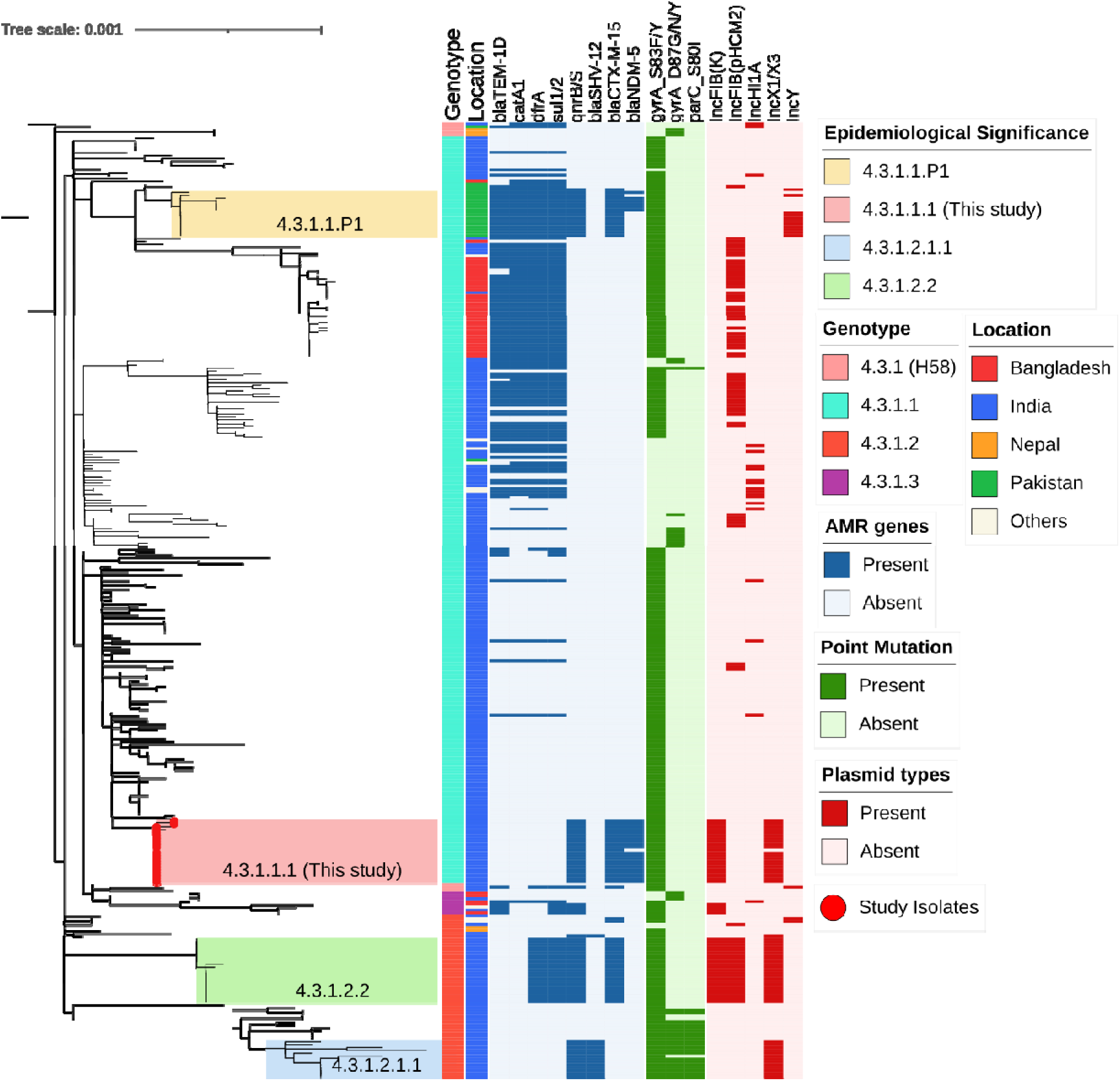
Phylogenetic relationship and genetic characteristics of 22 *S*. Typhi strains from India. A maximum likelihood phylogenetic tree was constructed using core genome SNPs from the study isolates (marked with red circles at the branch tips) along with global H58 isolates, revealing an overall SNP difference of 2,119 among the H58 isolates. Previously reported high-risk clones of cephalosporin-resistant clusters in the tree, including 4.3.1.2.1.1 (green), 4.3.1.1.P1 (blue), and 4.3.1.2.2 (orange). The colour-coded metadata strips adjacent to the tree represent: strip 1 denotes genotype, strip 2 indicates geographic region, and strip 3 shows the country location of each isolate. The heatmap alongside the tree depicts the distribution of antimicrobial resistance (AMR) genes, plasmid replicons, and point mutations, which are predominantly concentrated within the H58 cluster, reflecting its strong association with the spread of multidrug resistance.

### Characterization of Plasmids

All CRST isolates carried AMR genes located on both IncFIB(K) and IncX3 plasmids. AMR gene analysis revealed that the ∼ 73 kb IncFIB(K) plasmid harbored *bla*_CTX-M-15_, *qnrS1*, and *tet(A),* while the ∼ 47 kb IncX3 plasmid carried *bla*_NDM-5_ gene. To investigate the origin of these plasmids, complete circular sequences of IncFIB(K) and IncX3 plasmids from the study isolates were BLAST-compared with previously reported plasmids in the NCBI database. The IncFIB(K) plasmid (Accession no: CP189855) showed 100% sequence identity to an *E. coli* IncFIB(K) plasmid (CP116920), as well as to plasmids previously reported in ceftriaxone-resistant *S*. Typhi isolates from Gujarat, India (CP173298 and CP168964). However, the sequence coverage was 98% for *E. coli* and 93% for *S*. Typhi, indicating that while highly similar, the plasmids are not identical. Consistent with previous reports, *bla*_CTX-M-15_ in the IncFIB(K) plasmid were associated with mobilization by an IS1380 family ISEcp1 element located upstream (Figure 2a).

**Figure 2a:**
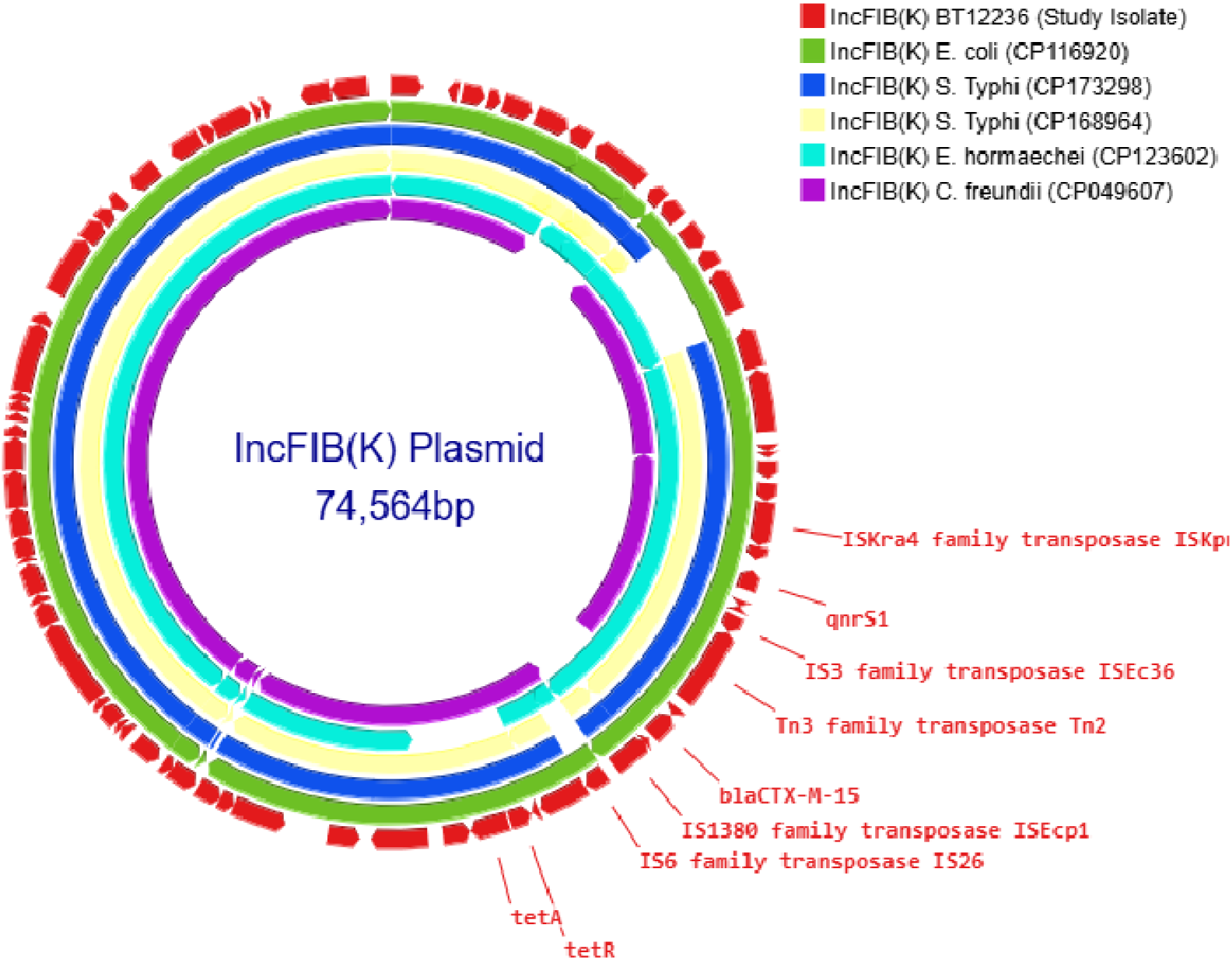
Comparison of IncFIB(K) plasmid: The IncFIB(K) plasmid from the carbapenem-resistant *Salmonella* Typhi (CRST) isolate was BLAST-searched against the NCBI BLASTn database and compared with Enterobacteriales carrying plasmids of the same incompatibility group [(CP116920), (CP173298), (CP168964), (CP123602), (CP049607)] with 90–100% identity. The selected plasmids were then annotated and visualized using Proksee.

A similar BLAST analysis of the IncX3 plasmid (Accession no: CP189856) carrying *bla*_NDM-5_ revealed 100% sequence identity and query coverage with IncX3 plasmids previously reported in *E. coli* (CP086557) and *K. pneumoniae* (CP080448). Notably, the IncX3 plasmid from this study showed only 83% query coverage with the SHV-carrying IncX3 plasmid (CP052768) identified from the *S*. Typhi outbreak in Mumbai, suggesting it is genetically distinct. The *bla*_NDM-5_ was located within the characteristic genetic structure ISAba125-IS5-*bla*_NDM–5_-*ble*_MBL_-trpF-dsbC-IS26 found in IncX3 type plasmids (Figure 2b).

**Figure 2b:**
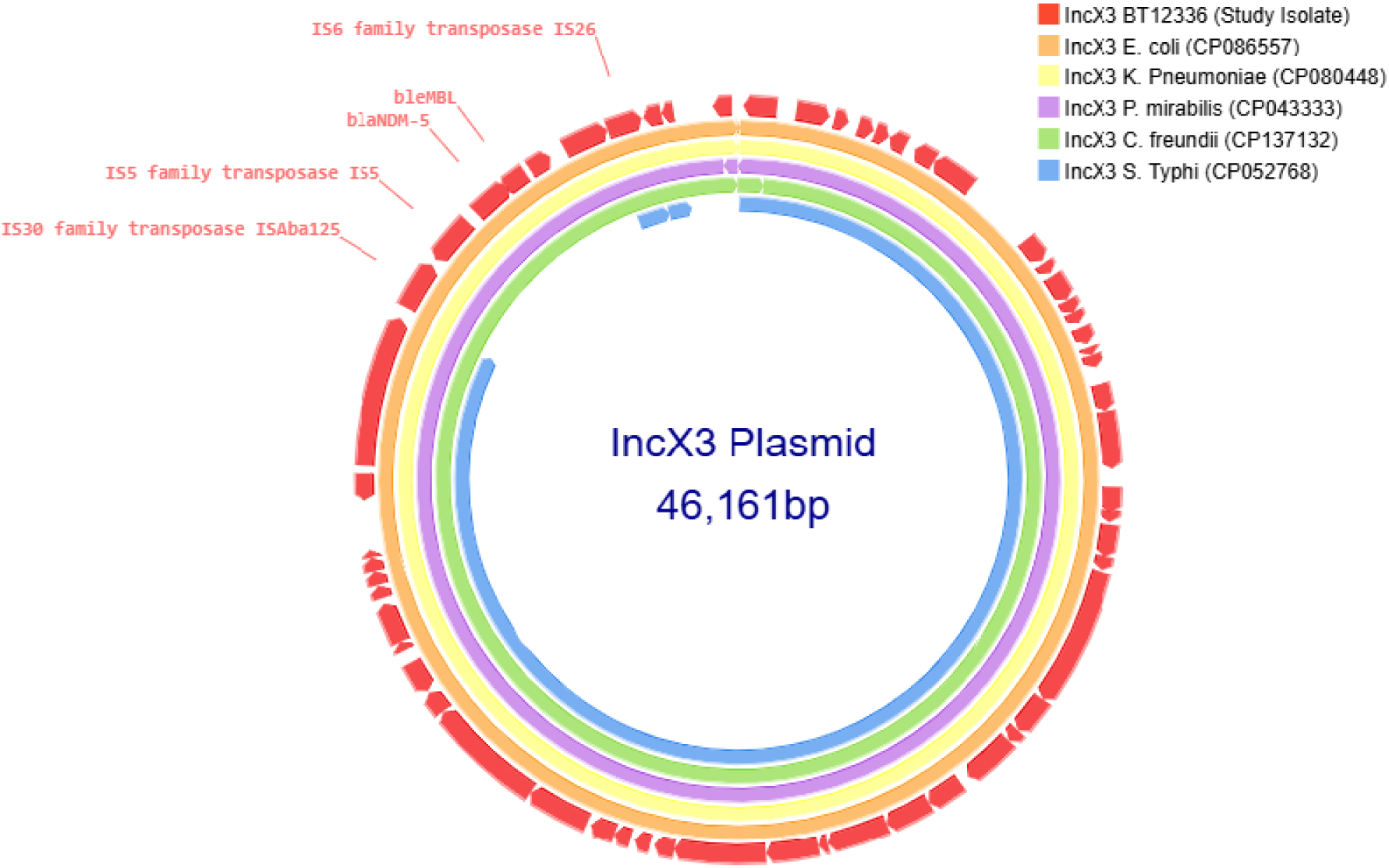
Comparison of IncX3 plasmid: The IncX3 plasmid from the carbapenem-resistant *Salmonella* Typhi (CRST) isolate was BLAST-searched against the NCBI BLASTn database and compared with Enterobacteriales carrying plasmids of the same incompatibility group [(CP086557), (CP080448), (CP043333), (CP137132), (CP052768)] with 90–100% identity. The selected plasmids were then annotated and visualized using Proksee.

## Discussion

The emergence of CRST signifies a critical evolutionary escalation, building on the shift from MDR to XDR strains observed in Pakistan by 2016 (Klemm et al., 2018). Before 2016, resistance in *S*. Typhi was primarily limited to first-line antibiotics and fluoroquinolones (FQNS), with the H58 lineage dominating across South Asia and Africa (Carey et al., 2024). Within this lineage, subclades 4.3.1.1 (MDR) and 4.3.1.2 (FQNS) were distinguishable via GenoTyphi genotyping (Wong et al., 2016). Subsequent emergence of ceftriaxone-resistant and XDR *S*. Typhi in India and Pakistan, respectively, was further mapped through finer subclade resolution (Dyson et al., 2021; Carey et al., 2023). For example, the Sindh XDR outbreak (4.3.1.1-P1) diverged by six SNPs from its nearest contemporaries (Klemm et al., 2018), while the Vadodara outbreak in India (4.3.1.2.1) exhibited 21 SNPs from its closest relative (Thirumoorthy et al., 2025). Notably, the CRST isolates in this study have accumulated seven unique chromosomal mutations (Table S3) while acquiring a *bla*_NDM-5_-harboring plasmid. Given the low mutation rate of *S*. Typhi (0.63 SNPs per genome per year; Wong et al.,2016), this shift appears to be a plasmid-driven phenotypic leap rather than gradual chromosomal adaptation, underscoring the role of horizontal gene transfer under selective antibiotic pressure (Rodríguez-Beltrán et al., 2021). These findings highlight unregulated antibiotic use in South Asia as a key driver of resistance evolution, emphasizing the urgent need for antimicrobial stewardship interventions.

The acquisition of diverse plasmids has played a pivotal role in the evolution of antimicrobial resistance in *S*. Typhi, driving the transition from MDR strains to emerging CRST variants. MDR *S*. Typhi strains harbored IncHI1 pST6 plasmids, almost exclusively within the H58 lineage, which became the dominant clade in South Asia and Africa (Holt et al., 2011). Resistance escalated further with the acquisition of *bla*_CTX-M-15_-carrying IncY plasmids, likely originating from *E.* coli, which facilitated the emergence of third-generation cephalosporin-resistant strains (Klemm et al., 2018). Subsequent ceftriaxone-resistant *S*. Typhi outbreaks in Mumbai and Vadodara, India, were linked to the horizontal acquisition of distinct plasmids, specifically IncX3 and IncFIB(K), respectively, likely sourced from co-circulating Enterobacteriaceae (Jacob et al., 2021; Thirumoorthy et al., 2025). This pattern indicates that *S*. Typhi has repeatedly leveraged plasmid pools from *E. coli* and *Klebsiella* spp., allowing it to bypass previously effective antibiotic therapies. The CRST isolates in this study represent the next stage in this evolutionary trajectory, having acquired an IncX3 plasmid harboring *bla*_NDM-5_ and an IncFIB(K) plasmid carrying *bla*_CTX-M-15,_ both likely derived from Enterobacteriaceae (Figure 2). Notably, the IncFIB(K) plasmid in the Vadodara outbreak differs from that found in CRST isolates, suggesting independent acquisition events rather than clonal dissemination. Similarly, the first reported carbapenem-resistant *S*. Typhi case in Peshawar, Pakistan (July 2022), carried *bla*_NDM-5_ on an IncN plasmid, again resembling plasmids found in other Enterobacteriaceae (Nizamuddin et al., 2023). The presence of multiple plasmid types (IncX3 and IncN) conferring carbapenem resistance suggests that CRST has arisen independently across different settings, rather than spreading from a single source. These findings highlight a rapidly evolving resistance landscape, where *S*. Typhi continues to integrate plasmid-borne resistance genes from Enterobacteriaceae, facilitating stepwise antibiotic resistance escalation.

In South Asia, MDR and FQNS *S*. Typhi infections are primarily treated with oral cefixime or azithromycin for outpatient cases, while intravenous ceftriaxone and azithromycin are used in combination for severe infections (Dolecek et al., 2019; Kuehn et al., 2022). For XDR or ceftriaxone resistant strains azithromycin remains the primary oral therapy for uncomplicated typhoid fever, whereas intravenous meropenem and azithromycin are recommended for severe cases (Qureshi et al., 2020; Parry et al., 2023). However, the emergence of CRST in India, co-producing *bla*_NDM_ and *bla*_CTX-M_ enzymes, renders both ceftriaxone and meropenem ineffective (Park et al., 2024). If CRST isolates remain susceptible to azithromycin (MIC ≤ 16 µg/mL), azithromycin remains the preferred oral treatment, leveraging its intracellular efficacy. Alarmingly, the first report of CRST from Peshawar, Pakistan, identified co-carriage of the *mphA* gene, conferring phenotypic resistance to azithromycin (Nizamuddin et al., 2023). In such cases, ceftazidime-avibactam + aztreonam combination therapy is recommended for severe infections (Tamma et al., 2021). Where aztreonam-avibactam is unavailable, colistin, fosfomycin, or tigecycline monotherapy may serve as alternative options (Parry et al., 2019). Given the limited clinical evidence for treating CRST, treatment decisions should be guided by individual patient factors, local resistance patterns, and expert consultation.

The rapid evolution of *S*. Typhi resistance, culminating in CRST, signals an urgent public health crisis in South Asia, where the failure of ceftriaxone and meropenem leaves severely limited treatment options for typhoid fever (Nabarro et al., 2022). Given the increasing ineffectiveness of antibiotics, preventive strategies must take priority to reduce disease burden and slow resistance evolution. One of the most effective interventions is the widespread adoption of the typhoid conjugate vaccine (TCV), which successfully curbed the Sindh XDR outbreak in Pakistan by reducing case numbers and lowering antibiotic selective pressure (Nampota-Nkomba et al., 2023; Qamar et al., 2024). Expanding TCV coverage across India and South Asia is critical to prevent CRST from becoming endemic (Mogasale et al., 2024). Beyond vaccination, strengthening water, sanitation, and hygiene (WASH) infrastructure is essential, as poor sanitation fuels *S*. Typhi’s fecal-oral transmission, sustaining high infection rates and increasing exposure to resistant strains (Luby et al., 2018). Enhanced genomic surveillance is also crucial for tracking CRST’s plasmid-driven spread and emerging resistance mutations—particularly *acrB*-R717Q/L mutations and *mphA*-mediated azithromycin resistance, as seen in Peshawar’s first CRST case (Nizamuddin et al., 2023). Finally, urgent antibiotic stewardship reforms are needed to curb the unregulated use of broad-spectrum antibiotics, a key driver of resistance escalation. Preserving azithromycin’s efficacy and ensuring restricted use of last-resort drugs like aztreonam-avibactam will be essential to maintaining effective treatment options for future cases.

In conclusion, the emergence of high-risk *S*. Typhi clones, particularly those harboring carbapenem resistance, underscores the urgent need for a comprehensive, multi-pronged strategy to contain their spread. Robust genomic surveillance is critical for tracking resistance trends, deciphering genetic evolution, and understanding transmission dynamics. Strengthening national antibiotic stewardship policies can help curb selective pressure and slow resistance escalation. Additionally, widespread typhoid conjugate vaccine (TCV) deployment can significantly reduce disease burden and limit antibiotic exposure. A coordinated global response, integrating surveillance, stewardship, and vaccination, is essential to mitigate the growing threat posed by carbapenem-resistant *S*. Typhi.

## Data Availability

All data produced in the present study are available upon reasonable request to the authors

## Acknowledgements

We gratefully acknowledge Drs. Duncan Steele and Supriya Kumar from the Bill & Melinda Gates Foundation for their technical guidance throughout the study, provided on behalf of the SEFI consortium. We extend our appreciation to all the members of the SEFI reference laboratory team at CMC Vellore for their dedicated efforts, with special thanks to Ms. Agila Kumari P, Ms. Baby Abirami S, Mr. Ayyanraj N, Ms. Sowmya Murugan, Ms. Yamini Umashankar, and Mr. Praveen Thilagan for their contributions to phenotypic testing and maintenance of stock cultures. We sincerely thank Dr. Anton Spadar (London School of Hygiene & Tropical Medicine, UK) for his significant contribution in updating the GenoTyphi genotyping scheme, enabling the assignment of new genotypes in this study.

The Bill & Melinda Gates Foundation (Investment ID INV-009497 OPP1159351) supported this study’s phenotypic work under the project “National Surveillance System for Enteric Fever in India.” G.K., B.V., and J.J. received support from the same grant. The genomic sequencing component was funded by the Indian Council of Medical Research (ICMR) through the Expression of Interest titled “To establish ICMR’s Genomic Surveillance for Antimicrobial Resistance” (Grant ID: AMR/DX/TYP/3/2024-CD). The funders had no role in the study design, data collection, management, analysis, interpretation, manuscript preparation, review, or the decision to submit the manuscript for publication.

## Ethical Approval

This study was approved by the Institutional Review Board (IRB) of Christian Medical College, Vellore (IRB Min No. 10393 dated 30.11.2016).

---

**Table S1:** Detailed patient demographics, clinical affiliations, and travel histories.

| S. No | Isolate ID | Clinical affiliations | Travel histories |
| --- | --- | --- | --- |
| 1 | ERR14867532 (B2288) | Indira Gandhi Institute of Child Health, Bangalore | No |
| 2 | ERR14867533 (B5777) | Baby Memorial Hospital, Kozhikode | NA |
| 3 | ERR14867544 (B5971) | Baby Memorial Hospital, Kozhikode | Yes |
| 4 | ADLM-01 | Agilus Diagnostic Labs Mumbai | NA |
| 5 | ERR14867534 (BT-15499) | St. John's Medical College Hospital, Bangalore | NA |
| 6 | ERR14867535 (BT-10862) | St. John's Medical College Hospital, Bangalore | NA |
| 7 | ERR14867536 (BT-15709) | St. John's Medical College Hospital, Bangalore | NA |
| 8 | ERR14867537 (BT-15401) | St. John's Medical College Hospital, Bangalore | NA |
| 9 | ERR14867538 (BT-14177) | St. John's Medical College Hospital, Bangalore | NA |
| 10 | ERR14867539 (BT-12236) | St. John's Medical College Hospital, Bangalore | NA |
| 11 | ERR14867540 (BT-17157) | St. John's Medical College Hospital, Bangalore | NA |
| 12 | ERR14867541 (BT-15985) | St. John's Medical College Hospital, Bangalore | NA |
| 13 | ERR1486754 (FB01) | Fortis Hospitals, Bengaluru | NA |
| 14 | ERR14867543 (PDH2) | P.D.Hinduja National Hospital & Medical Research Centre, Mumbai | NA |
| 15 | ERR14867545 (BC248) | KIMS, Trivandrum | Yes |
| 16 | ERR14867546 (BT-15515) | St. John's Medical College Hospital, Bangalore | NA |
| 17 | BA4920 | CMC, Chittoor Hospital | NA |
| 18 | 480/16319 | Fortis Hospitals, Bengaluru | NA |
| 19 | BT-1949 | St. John's Medical College Hospital, Bangalore | NA |
| 20 | 336/11763 | Fortis Hospitals, Bengaluru | NA |
| 21 | FB02 | Fortis Hospitals, Bengaluru | NA |
| 22 | BT-17019 | St. John's Medical College Hospital, Bangalore | NA |

**Supplementary Figure 1:**
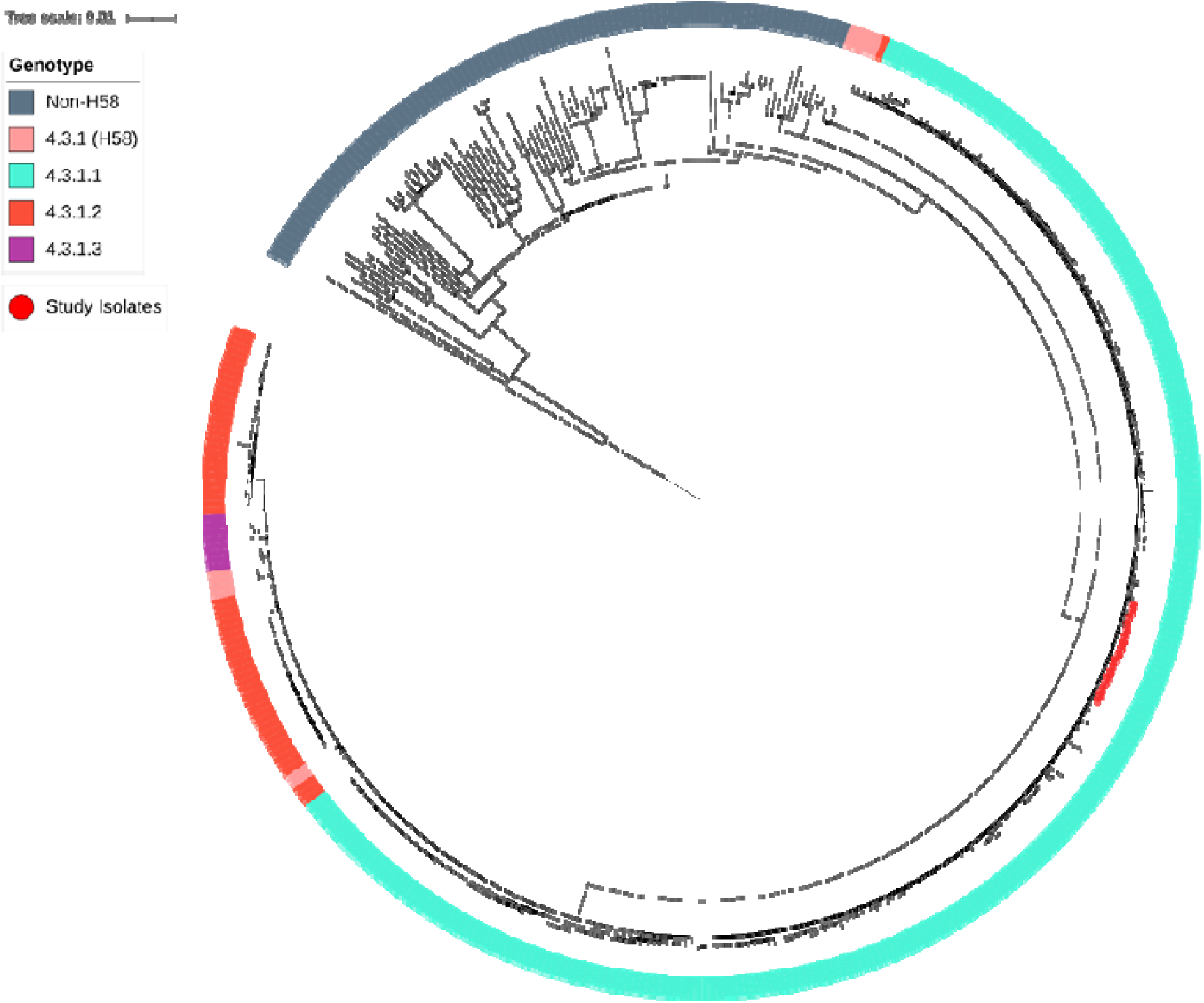
Phylogenetic relationship of 21 Carbapenem-resistant *S*. Typhi isolates from, India against 418 global *S*. Typhi strains. The maximum likelihood phylogenetic tree was constructed based on single nucleotide polymorphisms (SNPs) and mapped against the *Salmonella* Typhi CT18 reference strain, with an overall SNP count of 6,780. The genotypes of *S*. Typhi strains are represented as gradient-colored strips, while the study isolates are highlighted as red dots

## References

Parry CM, Hien TT, Dougan G, White NJ, Fever JF. 347. DOI: 10.1056/NEJMra020201. 2002:1770-82.

Crump JA, Sjölund-Karlsson M, Gordon MA, Parry CM. Epidemiology, clinical presentation, laboratory diagnosis, antimicrobial resistance, and antimicrobial management of invasive Salmonella infections. Clinical microbiology reviews. 2015 Oct;28(4):901–37.

Radhakrishnan A, Als D, Mintz ED, Crump JA, Stanaway J, Breiman RF, Bhutta ZA. Introductory article on global burden and epidemiology of typhoid fever. The American journal of tropical medicine and hygiene. 2018 Sep;99(3 Suppl):4.

Crump JA. Progress in typhoid fever epidemiology. Clinical Infectious Diseases. 2019 Feb 15;68(Supplement_1):S4-9.

Piovani D, Figlioli G, Nikolopoulos GK, Bonovas S. The global burden of enteric fever, 2017–2021: a systematic analysis from the global burden of disease study 2021. Eclinicalmedicine. 2024 Nov 1;77.

John J, Bavdekar A, Rongsen-Chandola T, Dutta S, Gupta M, Kanungo S, Sinha B, Srinivasan M, Shrivastava A, Bansal A, Singh A. Burden of typhoid and paratyphoid fever in India. New England Journal of Medicine. 2023 Apr 20;388(16):1491–500.

Browne AJ, Chipeta MG, Fell FJ, Haines-Woodhouse G, Hamadani BH, Kumaran EA, Aguilar GR, McManigal B, Andrews JR, Ashley EA, Audi A. Estimating the subnational prevalence of antimicrobial resistant Salmonella enterica serovars Typhi and Paratyphi A infections in 75 endemic countries, 1990–2019: a modelling study. The Lancet Global Health. 2024 Mar 1;12(3):e406-18.

Marchello CS, Carr SD, Crump JA. A systematic review on antimicrobial resistance among Salmonella Typhi worldwide. The American journal of tropical medicine and hygiene. 2020 Dec;103(6):2518.

Kirchhelle C, Dyson ZA, Dougan G. A biohistorical perspective of typhoid and antimicrobial resistance. Clinical Infectious Diseases. 2019 Oct 15;69(Supplement_5):S388-94.

Akram J, Khan AS, Khan HA, Gilani SA, Akram SJ, Ahmad FJ, Mehboob R. Extensively drug[resistant (XDR) typhoid: evolution, prevention, and its management. BioMed Research International. 2020;2020(1):6432580.

Klemm EJ, Shakoor S, Page AJ, Qamar FN, Judge K, Saeed DK, Wong VK, Dallman TJ, Nair S, Baker S, Shaheen G. Emergence of an extensively drug-resistant Salmonella enterica serovar Typhi clone harboring a promiscuous plasmid encoding resistance to fluoroquinolones and third-generation cephalosporins. MBio. 2018 Mar 7;9(1):10–128.

Jacob JJ, Pragasam AK, Vasudevan K, Veeraraghavan B, Kang G, John J, Nagvekar V, Mutreja A. Salmonella Typhi acquires diverse plasmids from other Enterobacteriaceae to develop cephalosporin resistance. Genomics. 2021 Jul 1;113(4):2171–6.

Thirumoorthy TP, Jacob JJ, Velmurugan A, Teekaraman MP, Shah B, Iyer V, Maheshwari G, Trivedi U, Shah A, Patel P, Gaigawale A. Recent emergence of cephalosporin-resistant Salmonella Typhi in India due to the endemic clone acquiring IncFIB (K) plasmid encoding bla CTX-M-15 gene. Microbiology Spectrum. 2025 Apr 10:e00875–24.

Carey ME, Jain R, Yousuf M, Maes M, Dyson ZA, Thu TN, Nguyen Thi Nguyen T, Ho Ngoc Dan T, Nhu Pham Nguyen Q, Mahindroo J, Thanh Pham D. Spontaneous emergence of azithromycin resistance in independent lineages of Salmonella Typhi in Northern India. Clinical Infectious Diseases. 2021 Mar 1;72(5):e120–7.

Baker S, Thomson N, Weill FX, Holt KE. Genomic insights into the emergence and spread of antimicrobial-resistant bacterial pathogens. Science. 2018 May 18;360(6390):733-8.

Wong VK, Baker S, Connor TR, Pickard D, Page AJ, Dave J, Murphy N, Holliman R, Sefton A, Millar M, Dyson ZA. An extended genotyping framework for Salmonella enterica serovar Typhi, the cause of human typhoid. Nature communications. 2016 Oct 5;7(1):12827.

Dyson ZA, Holt KE. Five years of GenoTyphi: updates to the global *Salmonella* Typhi genotyping framework. The Journal of infectious diseases. 2021 Dec 15;224(Supplement_7):S775-80.

Wong VK, Baker S, Pickard DJ, Parkhill J, Page AJ, Feasey NA, Kingsley RA, Thomson NR, Keane JA, Weill FX, Edwards DJ. Phylogeographical analysis of the dominant multidrug-resistant H58 clade of Salmonella Typhi identifies inter-and intracontinental transmission events. Nature genetics. 2015 Jun;47(6):632–9.

Pragasam AK, Pickard D, Wong V, Dougan G, Kang G, Thompson A, John J, Balaji V, Mutreja A. Phylogenetic analysis indicates a longer term presence of the globally distributed H58 haplotype of Salmonella Typhi in Southern India. Clinical Infectious Diseases. 2020 Oct 15;71(8):1856–63.

Carey ME, Thi Nguyen TN, Tran DH, Dyson ZA, Keane JA, Pham Thanh D, Mylona E, Nair S, Chattaway M, Baker S. The origins of haplotype 58 (H58) Salmonella enterica serovar Typhi. Communications Biology. 2024 Jun 28;7(1):775.

Carey ME, Dyson ZA, Ingle DJ, Amir A, Aworh MK, Chattaway MA, Chew KL, Crump JA, Feasey NA, Howden BP, Keddy KH. Global diversity and antimicrobial resistance of typhoid fever pathogens: Insights from a meta-analysis of 13,000 *Salmonella* Typhi genomes. Elife. 2023 Sep 12;12:e85867.

Argimón S, Nagaraj G, Shamanna V, Sravani D, Vasanth AK, Prasanna A, Poojary A, Bari AK, Underwood A, Kekre M, Baker S. Circulation of third-generation cephalosporin resistant Salmonella Typhi in Mumbai, India. Clinical Infectious Diseases. 2022 Jun 15;74(12):2234–7.

Ain Q, Tahir M, Sadaqat A, Ayub A, Awan AB, Wajid M, Ali A, Iqbal M, Haque A, Sarwar Y. First detection of extensively drug-resistant Salmonella typhi isolates harboring VIM and GES genes for carbapenem resistance from Faisalabad, Pakistan. Microbial Drug Resistance. 2022 Dec 1;28(12):1087–98.

Nizamuddin S, Khan EA, Chattaway MA, Godbole G. Case of carbapenem-resistant Salmonella Typhi infection, Pakistan, 2022. Emerging infectious diseases. 2023 Nov;29(11):2395.

Nair S, Patel V, Hickey T, Maguire C, Greig DR, Lee W, Godbole G, Grant K, Chattaway MA. Real-time PCR assay for differentiation of typhoidal and nontyphoidal Salmonella. Journal of clinical microbiology. 2019 Aug;57(8):10–128.

Performance Standards for Antimicrobial Susceptibility Testing Guideline M100, 34th ed.; Clinical and Laboratory Standards Institute: Malvern, PA, USA, 2024.

The European Committee on Antimicrobial Susceptibility Testing. Breakpoint Tables for Interpretation of MICs and Zone Diameters, Version 14.0. 2024. Available online: http://www.eucast.org/clinical_breakpoints

Poirel L, Walsh TR, Cuvillier V, Nordmann P. Multiplex PCR for detection of acquired carbapenemase genes. Diagnostic microbiology and infectious disease. 2011 May 1;70(1):119–23.

Grant JR, Enns E, Marinier E, Mandal A, Herman EK, Chen CY, Graham M, Van Domselaar G, Stothard P. Proksee: in-depth characterization and visualization of bacterial genomes. Nucleic acids research. 2023 Jul 5;51(W1):W484–92.

Croucher NJ, Page AJ, Connor TR, Delaney AJ, Keane JA, Bentley SD, Parkhill J, Harris SR. Rapid phylogenetic analysis of large samples of recombinant bacterial whole genome sequences using Gubbins. Nucleic acids research. 2015 Feb 18;43(3):e15-.

Page AJ, Taylor B, Delaney AJ, Soares J, Seemann T, Keane JA, Harris SR. SNP-sites: rapid efficient extraction of SNPs from multi-FASTA alignments. Microbial genomics. 2016 Apr 29;2(4):e000056.

Letunic I, Bork P. Interactive Tree Of Life (iTOL) v5: an online tool for phylogenetic tree display and annotation. Nucleic acids research. 2021 Jul 2;49(W1):W293–6.

Rodríguez-Beltrán J, DelaFuente J, León-Sampedro R, MacLean RC, San Millán Á. Beyond horizontal gene transfer: the role of plasmids in bacterial evolution. Nature Reviews Microbiology. 2021 Jun;19(6):347–59.

Holt KE, Phan MD, Baker S, Duy PT, Nga TV, Nair S, Turner AK, Walsh C, Fanning S, Farrell-Ward S, Dutta S. Emergence of a globally dominant IncHI1 plasmid type associated with multiple drug resistant typhoid. PLoS neglected tropical diseases. 2011 Jul 19;5(7):e1245.

Dolecek C, Pokharel S, Basnyat B, Olliaro P. Antibiotics for typhoid fever. In: The Selection and Use of Essential Medicines. WHO Technical Report Series 1021. Geneva: World Health Organization, 2019:19–26.

Kuehn R, Stoesser N, Eyre D, Darton TC, Basnyat B, Parry CM. Treatment of enteric fever (typhoid and paratyphoid fever) with cephalosporins. Cochrane Database of Systematic Reviews. 2022(11).

Qureshi S, Naveed AB, Yousafzai MT, Ahmad K, Ansari S, Lohana H, Mukhtar A, Qamar FN. Response of extensively drug resistant Salmonella Typhi to treatment with meropenem and azithromycin, in Pakistan. PLoS neglected tropical diseases. 2020 Oct 15;14(10):e0008682.

Parry CM, Qamar FN, Rijal S, McCann N, Baker S, Basnyat B. What should we be recommending for the treatment of enteric fever?. InOpen Forum Infectious Diseases 2023 May (Vol. 10, No. Supplement_1, pp. S26-S31). US: Oxford University Press.

Park SY, Baek YJ, Kim JH, Seong H, Kim B, Kim YC, Yoon JG, Heo N, Moon SM, Kim YA, Song JY. Guidelines for Antibacterial Treatment of Carbapenem-Resistant Enterobacterales Infections. Infection & Chemotherapy. 2024 Aug 2;56(3):308.

Tamma PD, Aitken SL, Bonomo RA, Mathers AJ, Van Duin D, Clancy CJ. Infectious Diseases Society of America guidance on the treatment of extended-spectrum β-lactamase producing Enterobacterales (ESBL-E), carbapenem-resistant Enterobacterales (CRE), and Pseudomonas aeruginosa with difficult-to-treat resistance (DTR-P. aeruginosa). Clinical Infectious Diseases. 2021 Apr 1;72(7):e169–83.

Parry CM, Ribeiro I, Walia K, Rupali P, Baker S, Basnyat B. Multidrug resistant enteric fever in South Asia: unmet medical needs and opportunities. Bmj. 2019 Jan 22;364.

Nabarro LE, McCann N, Herdman MT, Dugan C, Ladhani S, Patel D, Morris-Jones S, Balasegaram S, Heyderman RS, Brown M, Parry CM. British Infection Association guidelines for the diagnosis and management of enteric fever in England. Journal of Infection. 2022 Apr 1;84(4):469–89.

Nampota-Nkomba N, Carey ME, Jamka LP, Fecteau N, Neuzil KM. Using typhoid conjugate vaccines to prevent disease, promote health equity, and counter drug-resistant typhoid fever. InOpen Forum Infectious Diseases 2023 May (Vol. 10, No. Supplement_1, pp. S6-S12). US: Oxford University Press.

Qamar FN, Qureshi S, Haq Z, Yousafzai T, Qazi I, Irfan S, Iqbal N, Amalik Z, Hotwani A, Ali Q, Fatima I. Longevity of immune response after a single dose of typhoid conjugate vaccine against Salmonella Typhi among children in Hyderabad, Pakistan. International Journal of Infectious Diseases. 2024 Oct 1;147:107187.

Mogasale VV, Sinha A, John J, Farooqui HH, Ray A, Chantler T, Mogasale V, Dhoubhadel BG, Edmunds WJ, Clark A, Abbas K. Typhoid conjugate vaccine implementation in India: A review of supportive evidence. Vaccine: X. 2024 Oct 1:100568.

Luby SP, Rahman M, Arnold BF, Unicomb L, Ashraf S, Winch PJ, Stewart CP, Begum F, Hussain F, Benjamin-Chung J, Leontsini E. Effects of water quality, sanitation, handwashing, and nutritional interventions on diarrhoea and child growth in rural Bangladesh: a cluster randomised controlled trial. The Lancet Global Health. 2018 Mar 1;6(3):e302–15.

